# Functional Impacts of Aminoglycoside Treatment on Speech Perception and Extended High-Frequency Hearing Loss in Cystic Fibrosis

**DOI:** 10.1101/2020.04.29.20084848

**Authors:** Chelsea M. Blankenship, Lisa L. Hunter, M. Patrick Feeney, Madison Cox, Lindsey Bittinger, Angie Garinis, Li Lin, Gary McPhail, John P. Clancy

## Abstract

**Purpose:** The purpose of this study is to better understand the prevalence of ototoxicity-related hearing loss and its functional impact on communication in a pediatric and young adult cohort with cystic fibrosis (CF) and individuals without CF (controls).

**Method:** Observational, cross-sectional investigation of hearing function in children, teens, and young adults with CF (n = 57, mean = 15.0 yr.) who received intravenous aminoglycoside antibiotics and age- and gender-matched controls (n = 61, mean = 14.6 yr.). Participants completed standard and extended high frequency audiometry, middle ear measures, speech perception tests, and a hearing and balance questionnaire.

**Results:** Individuals with CF were 3 to 4 times more likely to report issues with hearing, balance, and tinnitus and performed significantly poorer on speech perception tasks compared to controls. A higher prevalence of hearing loss was observed in individuals with CF (57%) compared to controls (37%). CF and control groups had similar proportions of slight and mild hearing losses, however individuals with CF were 7.6 times more likely to have moderate and greater degrees of hearing loss. Older participants displayed higher average EHF thresholds, with no effect of age on average SF thresholds. Although middle ear dysfunction has not previously been reported to be more prevalent in CF, this study showed that 16% had conductive or mixed hearing loss and higher rates of previous otitis media and pressure equalization (PE) tube surgeries compared to controls.

**Conclusions:** Individuals with CF have a higher prevalence of conductive, mixed and sensorineural hearing loss, poorer speech-in-noise performance and higher rates of multiple symptoms associated with otologic disorders (tinnitus, hearing difficulty, dizziness, imbalance and otitis media) compared to controls. Accordingly, children with CF should be asked about these symptoms, receive baseline hearing assessment(s) prior to treatment with potentially ototoxic medications, and at regular intervals thereafter in order to provide otologic and audiologic treatment for hearing and ear-related problems to improve communication functioning.

## Introduction

Cystic fibrosis (CF) is the most common congenital life-threatening genetic disease in Caucasians occurring in 1/2500 births and affecting over 70,000 individuals worldwide (Cystic Fibrosis Foundation, 2017; Kreicher, Bauschard, Clemmens, Riva, & Meyer, 2018). Individuals with CF experience recurrent pulmonary infections that are commonly treated with intravenous aminoglycoside (IV-AG) and glycopeptide antibiotics (Barclay et al., 1996; Becker & Cooper, 2013; Jiang, Karasawa, & Steyger, 2017; O’Sullivan et al., 2017). Due in part to the aggressive IV-AG treatment, the predicted median age of survival for patients with CF has increased from 28 years in 1989 to 43 years in 2016 with the number of survivors over 40 years of age doubling every 5 years (Cystic Fibrosis Foundation, 2017). Individuals with CF often require repeated doses of IV-AG treatment throughout life, and thus are at a higher risk of developing permanent sensorineural hearing loss (SNHL; cochleotoxicity) and balance disorders (vestibulotoxicity; Ariano et al., 2008; Garinis et al., 2017; Handelsman et al., 2017; Tan et al., 2003). In some cases, the effects of ototoxicity can be severe and life altering. The stark reality of going deaf due to aminoglycoside (AG) antibiotics was poignantly written in the words of Brad Dell, a 25-year-old man with cystic fibrosis, “I’ll never forget the day of nothing. Hearing absolutely nothing. The remainder of my hearing had leaked out of my ears as more amikacin doses leaked into my veins” (Dell, 2018). As hearing loss progresses, speech perception is subsequently affected, highlighting the importance of speech perception assessment in ototoxicity monitoring programs.

Aminoglycoside (e.g., amikacin, tobramycin and gentamicin) and glycopeptide (e.g., vancomycin) antibiotics remain the primary treatment agent of choice for CF pulmonary exacerbations and are in widespread use in CF centers (Barclay et al., 1996; Becker & Cooper, 2013; Jiang et al., 2017; O’Sullivan et al., 2017). They are cost-effective and highly efficient against gram-negative organisms, such as *Pseudomonas aerugonisa*, which are the underlying cause of pulmonary lung infections in CF (Conway, Miller, Ramsden, & Littlewood, 1985; Szaff, Hoiby, & Flensborg, 1983). The median age at which *Pseudomonas aerugonisa* infections first occurs is 5.2 years. Using the Leeds classification criteria, 46.6% of all individuals with CF have *Pseudomonas aerugonisa*, while 28.6% are chronic and necessitate repeated courses of IV-AG treatment (Cystic Fibrosis Foundation, 2017). While treatment with AGs alone or in conjunction with glycopeptides is common practice to treat life-threatening bacterial infections, these antibiotics have a well-documented ototoxic effect, resulting in irreversible damage to hearing and balance structures within the inner ear (Handelsman, Nasr, Pitts, & King, 2017; Liu et al., 2013; Pauna, Monsanto, Kurata, Paparella, & Cureoglu, 2017; Sone, Schachern, & Paparella, 1998). Ototoxicity can result as a direct effect of AGs, with gentamicin considered to be primarily vestibulotoxic, and amikacin and tobramycin mainly cochleotoxic. However, in theory they can all induce damage to both parts of in the inner ear (Jiang et al., 2017; Selimoglu, 2007). Glycopeptide antibiotics (e.g., vancomycin) can indirectly cause hearing loss due to their synergistic effect with AGs. For example, vancomycin is highly nephrotoxic, resulting in decreased kidney function/renal clearance of AGs potentially leading to an increased susceptibility to developing hearing loss (Filippone, Kraft, & Farber, 2017).

The cochleotoxic effect of aminoglycosides, with or without glycopeptides, is characterized by SNHL that originates in the basal turn of the cochlea. Within the high frequency region, outer hair cell loss occurs first followed by inner hair cell loss after which damage progresses to the apical region affecting lower frequencies (Fausti et al., 1984; Guthrie, 2008; Huizing & de Groot, 1987). Thus, extended high frequency (EHF; > 8 kHz) audiometry is integral to ototoxicity monitoring due to the progression of hearing loss from high to low frequencies (Fausti, Frey, Henry, Olson, & Schaffer, 1993; Fausti et al., 1992; Fausti et al., 1994). Furthermore, Fausti et al. (1999) identified a sensitive (frequency) range for ototoxicity (SRO) which focuses on the highest frequencies that are audible to an individual patient. The SRO method involves defining the highest audible frequency and then measuring pure-tone thresholds in one-sixth octaves up to the predetermined frequency limit. This shortened ototoxicity monitoring test decreases overall test time to approximately 10 minutes and maintains a 90% detection rate of significant changes in hearing (Fausti et al., 2003; Fausti et al., 1999).

The prevalence of hearing loss in children with CF repeatedly treated with IV-AGs ranges from 0 to 44% (Al-Malky, Dawson, Sirimanna, Bagkeris, & Suri, 2015; Thomsen, Friis, Jensen, Bak-Pedersen, & Larsen, 1979) and extends up to 59% in adults (Garinis et al., 2017; Scheenstra, Heijerman, Zuur, Touw, & Rijntjes, 2010; Zettner & Gleser, 2018). Substantial variability in prevalence rates occurs across studies due to differences in the population studied, type of AG and potential co-administration of glycopeptides, mode of drug administration, total body clearance of the drug, total number of AG doses, individual susceptibility, audiologic test protocol and hearing loss criteria. In general studies that only included the standard frequencies (SF; 0.25 to 8 kHz) found lower rates of hearing loss (Cheng et al., 2009; Forman-Franco, Abramson, Gorvoy, & Stein, 1979; Ozcelik et al., 1996; Stavroulaki et al., 2002) compared to studies that included EHF audiometry (Al-Malky et al., 2015;Al-Malky, Suri, Dawson, Sirimanna, & Kemp, 2011; Garinis et al., 2017; Martins, Camargos, Becker, Becker, & Guimaraes, 2010; Pedersen, Jensen, Osterhammel, & Osterhammel, 1987). In pediatric patients, Al-Malky et al. (2015) and Cheng et al. (2009) reported a 3 to 24% prevalence of SNHL with less than 10 doses of IV-AGs that increased to 40-44% with recurrent AG exposure (> 10 doses). In mostly adults with CF, individuals with higher cumulative doses of IV-AGs were 4.5 times more likely to have hearing loss compared to individuals with lower cumulative doses (Garinis et al., 2017). In a recent retrospective examination of the prevalence and type of hearing loss in the pediatric CF population, 31.8% had hearing loss in at least one ear in the SF region (Kreicher et al., 2018). Further analysis of the type of hearing loss (considering each ear separately) showed that 74.1% had normal hearing, 5.3% had conductive loss, 2.3% sensorineural loss, 5.1% mixed hearing loss, and 13.2% undefined (Kreicher et al., 2018).

Even though the use of EHF audiometry was first reported in the 1980s (Ahonen & McDermott, 1984; Fausti et al., 1984; McRorie, Bosso, & Randolph, 1989), nearly 40 years later it is still not routinely used in audiology clinics to monitor changes in hearing due to ototoxicity. Potential explanations include lack of clear ototoxicity monitoring guidelines and protocols, time constraints, audiology equipment and staffing limitations, absence of EHF audiometry reimbursement codes, as well as outcome data that indicate preserving EHF is important and has a positive impact on overall qualify of life. A survey of Cystic Fibrosis Foundation Accredited Care Centers and Affiliated Programs reported that while 43% of clinics performed audiological evaluations, it is unclear if EHF were included and some centers only measured thresholds if the patient was symptomatic (Van Meter, Corriveau, Ahern, & Lahiri, 2009). More recently, Prescott (2011) reported that 39% of pediatric CF programs do not monitor for ototoxicity at all, 32% of clinics monitor hearing in the SF range, and 46% include EHF audiometry. In addition, less than half of published pediatric studies of AG associated ototoxicity include EHF audiometry. This likely because most pediatric CF studies are retrospective. Since EHF audiometry is not routinely used in clinical practice, the onset and true prevalence of ototoxic hearing loss is likely underestimated.

In contrast to the common misconception that EHFs contribute very little to speech understanding, substantial information occurs at frequencies above 8 kHz that can be used for localization, speaker identification, and phoneme identification. Alexander, Kopun, and Stelmachowicz (2014) reported fricatives /ch, f, j, s, sh, th, v, and z/ contain differential spectral information from 3 to 10 kHz that can be used for phoneme identification. Furthermore, EHF spectral information aides in vowel and consonant identification when access to SF energy is restricted or degraded (Lippmann, 1996; Vitela, Monson, & Lotto, 2015) or when extended bandwidth hearing aids improve audibility of EHFs (Seeto & Searchfield, 2018). EHF improves sound localization by providing cues to resolve front/back confusions (Best, Carlile, Jin, & van Schaik, 2005; Brungart & Simpson, 2009; Heffner & Heffner, 2008) and can help listeners identify the speaker and segregate their voice from background talkers (Monson, Rock, Schulz, Hoffman, & Buss, 2019). In adults, Yeend, Beach, and Sharma (2019) reported poorer EHF thresholds are significantly correlated with poorer speech understanding in noise on the Listening in Spatialized Noise Task high-cue condition (Cameron & Dillon, 2007), and increased self-report of listening difficulties (Speech, Spatial and Qualities of Hearing; (Gatehouse & Noble, 2004). Additionally, Motlagh Zadeh et al. (2019) reported that adults with self-reported difficulty following conversations in noise were more likely to have EHF hearing loss (>20 dB HL).

Early detection of hearing loss is extremely important in pediatric patients due to its documented impact on speech and language development, literacy development (Yoshinaga-Itano, 1999, 2003), and scholastic achievement (Moeller, Tomblin, Yoshinaga-Itano, Connor, & Jerger, 2007). Children and teens with CF that receive greater or more frequent doses of ototoxic antibiotics will likely experience EHF hearing loss that accumulates over time into adulthood. (Al-Malky et al., 2015; Al-Malky et al., 2011; Garinis et al., 2017). As AG ototoxicity damage progresses from base to apex, frequencies in the human speech range will ultimately be involved, causing a permanent communication disability. Although EHF hearing loss is assumed to be associated with progressive declines in speech perception (Besser, Festen, Goverts, Kramer, & Pichora-Fuller, 2015; Hunter et al., 2020b; Levy, Freed, Nilsson, Moore, & Puria, 2015; Motlagh Zadeh et al., 2019), there is little if any published data on speech-in-noise performance in individuals with CF and AG exposure. Furthermore, the functional impact of speech understanding in noise is highly relevant for environments that children and adults listen in, such as the classroom, workplace, and social settings. Other notable limitations of previous studies include absence of EHF audiometry or bone-conduction (BC) thresholds, which limits their ability to accurately identify the presence and classify type of hearing loss. Most studies did not include an age-matched control group to make a direct and accurate comparison to the general population. Lastly, as previously mentioned, none included a functional measure of hearing loss on daily communication or evaluation of patient and parent perception of perceived hearing difficulties. These patient-reported outcomes are critical for improving patient-centered clinical care approaches and for improving communication between the patient and their clinical care team.

The goal of this study was to better understand the prevalence of ototoxicity-related hearing loss and its functional impact on communication in a pediatric cohort with CF. In order to achieve this goal, we examined SF and EHF audiometry (air- and bone-conduction), tympanometry, and middle ear muscle reflexes (MEMR) to fully describe hearing function in children with CF children and age- and gender-matched controls. A standardized pediatric measure of speech-in-noise as well as a self-report questionnaire for patients and parents on perceived hearing and balance difficulties and the functional impact of hearing loss on communication was also completed.

## Materials and Methods

### Participants

The study was part of a larger longitudinal project to evaluate the prevalence of hearing loss in children and adults with CF in relation to the concentration of AG-drug exposure and toxicologic response (pharmacodynamics), including ototoxic outcomes. Children and young adults diagnosed with CF (n = 57, mean age = 15.0 yr., range = 6.0 to 21.0) who received tobramycin or amikacin IV antibiotics, alone or in combination with vancomycin were recruited from the CF inpatient unit at Cincinnati Children’s Hospital Medical Center (CCHMC). Age- and gender-matched controls (n = 61, mean age = 14.6 yr., range = 7.5 to 21.1) were recruited from the same hospital through internal staff emails and outpatient study flyers. Additional eligibility criteria for both the CF and control group included ages between 6-21 years of age and the ability to complete a conventional behavioral hearing test. Controls were not excluded based on hearing status in order to provide a more accurate representation of the hearing levels of the general population. They had no history of AG treatment, and no CF diagnoses. The study was approved by the CCHMC Institutional Review Board. Informed parental consent and child assent (for ages 9-17 years) or patient consent (ages 18 and older) was obtained prior to enrollment. All participants were reimbursed for their participation.

### Test Protocol

All participants completed comprehensive diagnostic assessments including otoscopy, 226-Hz tympanometry, MEMRs, SF and EHF audiometric assessment, and speech understanding in quiet and noise. A hearing and balance questionnaire was administered to determine if the participant or the parent reported any hearing difficulties, tinnitus, balance disturbance, history of otitis media, PE tubes, or previous hearing exams. Additional details regarding onset, frequency and type of symptoms, and family history of hearing loss were also queried (see questionnaire in Appendix 1). This was a non-validated questionnaire, comparable to a case history or intake questionnaire that would be filled out during a clinical audiology appointment. All test procedures were completed in a double-walled sound booth (Industrial Acoustics Company, North Aurora, Illinois). Otoscopy was completed first to ensure the ear canal was patent, and if necessary, cerumen was removed. The test protocol took approximately one hour to complete. Individuals with CF completed testing while they were inpatient at CCHMC, while controls were tested during an outpatient research appointment.

<u>Audiometric Assessment:</u> Air-conduction (AC) thresholds were obtained with an Equinox Audiometer (Interacoustics Inc., Middlefart, Denmark) and Sennheiser HDA-300 circumaural headphones (Old Lyme, CT) in the SF range at octave test frequencies from 0.25 to 8 and in the EHF range (10, 12.5, 14 and 16 kHz). Calibration was completed according to ISO 389.8 (2004) for SF and ISO 389-1 (2017) for EHF. Bone-conduction (BC) thresholds were measured using a Radio-ear B-71 bone oscillator (Radioear Corp, New Eagle, PA) at 0.25, 0.5, 1, 2, and 4 kHz if AC thresholds were ≥ 20 dB HL with appropriate narrowband masking in the contralateral ear. A modified Hughson-Westlake procedure with a 5-dB step size was used to measure threshold. For both the SF and EHF range, audiometric thresholds were used to categorize each ear based on the presence, degree, and type of hearing loss as well as the ear affected (left, right, bilateral). The degree of hearing loss was classified according to Goodman (1965) and Northern and Downs (1984) criteria for the pure-tone average for 0.5, 1, and 2 kHz: normal (0 to 15 dB HL), slight (16-25 dB HL), mild (26-40 dB HL), moderate (41-55 dB HL), moderately-severe (56-70 dB HL), severe (71-90 dB HL), and profound (91 + dB HL). The type of hearing loss was classified as follows:

1. <u>Normal Hearing (NH):</u> AC and BC thresholds within the normal range (≤ 15 dB HL).
2. <u>Conductive Hearing Loss (CHL):</u> BC thresholds less than or equal to 15 dB HL, air-conduction thresholds greater than 15 dB, and an air-bone gap of 10 dB at two frequencies or 20 dB at one frequency. For some participants abnormal tympanometry values were used to diagnose conductive loss (see criteria below in 226-Hz Tympanometry description).
3. <u>Sensorineural Hearing Loss (SNHL):</u> AC and BC thresholds greater than 15 dB HL without an air-bone gap (defined under CHL) at any test frequency.
4. <u>Mixed Hearing Loss (MHL):</u> Presence of a sensorineural and conductive hearing loss at the same audiometric test frequency or within the same ear.

<u>Middle Ear Measures:</u> Middle ear function was measured using a traditional 226-Hz tympanometry using the Titan (Interacoustics Inc., Middlefart, Denmark). Individuals were not excluded due to middle ear dysfunction. If BC thresholds were not measured (due to time restraints or patient fatigue), tympanometry values were used to determine if there was a conductive component to the hearing loss with the following normative ranges for tympanometric peak pressure (−100 to +30 daPa), admittance (0.3 to 1.5 mmho) and tympanometric width (30 to 105 daPa; Roup et al., 1998). At least two out of three values needed to be identified as abnormal (i.e., outside normative range) for the individual to be classified as having a conductive component to the hearing loss.

<u>Middle Ear Muscle Reflexes (MEMR).</u> MEMRs were assessed using the Titan (Interacoustics Inc., Middlefart, Denmark) clinical system alone, or in combination with a research wideband absorbance technique with custom designed recording system. MEMR thresholds were measured using a broadband noise stimulus in ipsilateral (same ear) and contralateral (opposite ear) conditions. In both systems, ear canal air pressure was adjusted to the peak tympanometric pressure obtained during wideband tympanometry (results not reported here). For the Interacoustics Titan clinical system, the broadband stimuli (0.5 to 8 kHz) were presented from 60 to 100 dB SPL in 5 dB steps. An ascending technique was used where the lowest intensity that evoked a reflex using a “very sensitive” criteria of 0.02 ml was marked as threshold. With the research wideband absorbance technique, MEMRs were measured using a pulsed-activator stimulus set that included four broadband noise activator (0.25 to 8 kHz) pulses that alternated with five wideband (0.25 to 8 kHz) clicks. Stimulus levels were calibrated in a 2-cm (HA-1) coupler and were presented in 5-dB steps from 60 to 120 dB peSPL. MEMRs measured with the research wideband absorbance technique used response averaging, artifact rejection and signal processing techniques to measure threshold, onset latency and amplitude growth. Additional details regarding the measurement and analysis procedures may be found in Keefe, Feeney, Hunter, and Fitzpatrick (2017). The control group had relatively equal percentages of MEMRs measured using the Titan (52%) compared to the wideband research technique (48%). In comparison, the CF group had a higher percentage of MEMRs measured with the wideband research technique (68%) compared to the Titan (32%). Since the wideband technique results in lower MEMR thresholds, a correction factor was applied in order to combine MEMR measured using the two different recording systems. The correction factor was based on MEMR thresholds measured from a group of children from a separate study using an identical MEMR test protocol (Hunter et al., 2020a). The children all had typical development and normal hearing in the SF region. That normative sample included 48 children with typical development (93 ears) with normal hearing that ranged in age from 6.5 to 14.5 (mean = 10.0 yr.). The correction factor applied to the research-based thresholds was 15.5 dB and 12.5 dB SPL for the ipsilateral and contralateral MEMR reflexes, respectively.

<u>Speech Perception Testing.</u> A speech reception threshold (SRT) was measured for each ear, using recorded spondees from the Central Institute for the Deaf W-1 adult or child word list (Auditec, 2015; Hirsch et al., 1952). Speech understanding in noise was evaluated using the Bamford-Kowal-Bench Speech-in-Noise (BKB-SIN) test measured at 50 dB HL in the monaural condition with 1 list pair administered to each ear (Etymōtic Research, 2005). This is an adaptive measure with signal-to-noise ratio (SNR) values ranging from +21 to -6 dB and is representative of environments that occur in the classroom and everyday settings. The participants were instructed to repeat each sentence that they heard and to guess if they were unsure. The number of key words repeated correctly for each sentence was tallied and used to calculate the SNR in dB necessary for the individual to understand 50% of the sentence (SNR-50). The SNR-Loss was calculated as the individual SNR-50 minus age-matched normative SNR-50 values from the BKB-SIN manual. The SNR-Loss is used to determine how much greater a SNR is needed for the individual to have equivalent performance to their age-matched peers. Additionally, the SNR-Loss can be categorized as follows to indicate the degree of difficulty understanding speech-in-noise: 1.) Normal is 0 to 3 dB, 2.) Mild is greater than 3 to 7 dB, 3.) Moderate is greater than 7 to 15 dB, 4.) Severe is greater than 15 dB (Etymōtic Research, 2005).

### Statistical Analysis

Descriptive statistics were used to summarize demographics and outcome measurements to identify any errors and outliers. Interval variables were summarized by central tendency and dispersion, and categorical variables were described by frequencies and percentages. Two-sample t-test, Chi-Square test or Fisher Exact tests were performed as appropriate to compare the demographics between the CF and control groups. Boxplots were created to study the distribution of the outcome variables. Mixed models were conducted to study the differences between the CF and control groups, with Test Ear included as a repeated measure and demographic factors included as covariates (Age at Test, Sex, and Race). Where appropriate, Test Frequency was included as a repeated measure and Hearing Status was included as a fixed effect factor. Interaction terms were also studied, and in some cases were removed from the final adjusted model for a parsimonious model due to their insignificance (*p* > 0.05). Sidak adjustment was applied for pairwise comparisons among the significant factors. Pearson correlations were used to assess the relationship between audiometric thresholds, SNR-Loss, and Age at Test. All data were collected and managed using REDCap, which is a secure web-based software platform (Harris et al., 2019; Harris et al., 2009) and then exported and formatted for analysis using SAS statistical software version 9.3 (SAS Institute, Cary, N.C.). Two-sided significance level was set at 0.05 for all analyses.

## Results

### Cohort Characteristics

A total of 74 individuals with CF were approached for the study, however 10 individuals were not interested in participating (14%) and 7 individuals were consented but were unable to complete testing prior to discharge (9%) resulting in 57 individuals that were enrolled and completed test procedures (77%). A control group of children without CF (control; n = 61) were enrolled and completed testing. Therefore, the study included 118 participants, 57 individuals with CF (49%), and 61 controls (51%). For individuals with CF, the mean age at test was 15.0 years (SD = 3.6, range = 6.0 to 21.0), 42% were males and 98% were Caucasian. In the control group, the mean age at test was 14.6 years (SD = 2.9, range = 7.5 to 21.1), 46% were males, and 85% were Caucasian. There were no significant group differences in Age at Test and Sex however the control group had a higher number of non-Caucasians than the CF group. See Table 1 for additional group characteristics.

**Table 1.** Study sample demographics, questionnaire, and hearing loss characteristics for all participants combined, as well as the CF and control groups.

|  | All | CF | Control | *p*-value |
| --- | --- | --- | --- | --- |
| **Number of Participants** | 118 | 57 | 61 |  |
| **Age at Test** |  |  |  | 0.534‡ |
| Mean (SD) | 14.77 (3.28) | 14.96 (3.63) | 14.59 (2.94) |  |
| Range | 6.0-21.1 | 6.0-21.0 | 7.5-21.1 |  |
| **Sex\*** |  |  |  | 0.678# |
| Male | 52 (44.07) | 24 (42.11) | 28 (45.90) |  |
| Female | 66 (55.93) | 33 (57.89) | 33 (54.10) |  |
| **Race\*** |  |  |  | 0.017^ |
| Caucasian | 108 (91.53) | 56 (98.25) | 52 (85.25) |  |
| Non-Caucasian | 10 (8.47) | 1 (1.75) | 9 (14.75) |  |
| **Hearing and Balance Questionnaire\*** |  |  |  |  |
| Hearing Problem | 23 (19.49) | 18 (31.57) | 5 (8.19) | 0.001# |
| Tinnitus | 39 (33.05) | 30 (52.63) | 9 (16.07) | <0.001# |
| Balance Issues | 23 (19.49) | 18 (31.57) | 5 (8.92) | 0.003# |
| History of Otitis Media | 40 (33.89) | 24 (42.10) | 16 (26.22) | 0.049# |
| PE Tubes | 16 (13.55) | 9 (15.78) | 7 (11.47) | 0.412# |
| Previous Hearing Test | 97 (82.20) | 47 (82.45) | 50 (89.28) | 0.919# |
| **Hearing Status†** |  |  |  |  |
| Normal | 126 (53.39) | 49 (42.98) | 77 (63.11) | — |
| Hearing Loss | 110 (46.61) | 65 (57.02) | 41 (36.89) | — |
| **Hearing Loss Frequency Range†** |  |  |  |  |
| Standard Frequency | 64 (27.59) | 43 (37.72) | 21 (17.21) | — |
| Extended High Frequency | 91 (39.22) | 54 (47.37) | 37 (30.33) | — |
| **Degree of Hearing Loss†** |  |  |  |  |
| Slight | 53 (22.46) | 24 (21.05) | 29 (23.77) | — |
| Mild | 27 (11.44) | 15 (13.16) | 12 (9.84) | — |
| Moderate | 21 (8.09) | 18 (15.79) | 3 (2.46) | — |
| Moderate-Severe | 6 (2.54) | 5 (4.39) | 1 (0.82) | — |
| Severe | 3 (1.27) | 3 (2.63) | 0 (0.00) | — |
| **Type of Hearing Loss†** |  |  |  |  |
| Conductive | 8 (3.39) | 6 (5.26) | 2 (1.64) | — |
| Sensorineural | 82 (34.75) | 47 (41.23) | 35 (28.69) | — |
| Mixed | 18 (7.63) | 12 (10.53) | 6 (4.92) | — |
| Undetermined | 2 (0.85) | 0 (0.00) | 2 (1.64) | — |

Note: PE = Pressure Equalizer Tube, \* = number (%) of participants, † = number (%) of ears, ‡ = Two-Sample t-test, # = Chi-Square test, ^ Fisher's Exact Test

### Hearing and Balance Questionnaire

Parent and participant hearing and balance questionnaire responses were combined to determine an overall response for each question for each participant. Questions regarding hearing concerns, tinnitus, and balance issues were binary (Yes/No), while history of otitis media, PE tubes and previous hearing test were categorical (Yes/No/Not sure). Additional details regarding onset, frequency and type of symptoms were also queried (see questionnaire in Appendix 1). Table 1 displays the number of individuals within the CF and control group and corresponding percentages that indicated “Yes” to symptoms included in the questionnaire. Chi-square tests were performed to examine differences in hearing and balance concerns between the CF and control group. Individuals with CF reported significantly more hearing concerns (CF = 32%, Control = 8%), tinnitus (CF = 53%, Control = 16%), and balance problems including dizziness, unsteadiness or imbalance (CF = 32%, Control = 9%) compared to controls. Individuals with CF were also more likely to have a history of middle ear infections than controls (CF = 42%, Control = 26%). Although non-significant, the CF group reported a slightly higher history of PE tubes (CF = 16%, Control = 11%) with controls reporting a higher percentage of having a previous hearing test (CF = 82%, Control = 89%).

### Audiometric Thresholds

Descriptive statistics for prevalence, degree and type of hearing loss are displayed in Table 1. Individuals with CF had a significantly higher prevalence of hearing loss in either the SF or EHF ranges (57%) compared to controls (37%). Within the CF group, 38% had hearing loss in the SF range; the majority (88%) had slight to mild hearing loss while a smaller percentage (12%) had moderate to moderate-severe hearing loss. In the EHF range, 47% of individuals with CF had hearing loss, with almost equal percentages of slight to mild hearing loss (52%) and moderate to severe hearing loss (48%). In the control group, only 37% had hearing loss; 17% of control ears had hearing loss in the SF range and 30% had hearing loss in the EHF range. Of the control ears with hearing loss in the SFs, the degree of loss only ranged from slight to mild with no greater degrees of hearing loss present. In the EHF range, most controls had hearing loss in the slight to mild range (89%), with few ears in the moderate to moderately severe range (11%). When examining the degree of hearing loss across all audiometric frequencies, the CF and control group have similar numbers of slight and mild hearing losses, however individuals with CF had much greater percentages of moderate and greater degrees of hearing loss (CF = 23%, Control = 3%). With regard to the type of hearing loss, 5% of the CF group had a CHL, 41% had SNHL, and 11% had MHL. Controls mainly had SNHL (29%), with smaller percentages of CHL (2%) and MHL (5%).

The CF and control group mean audiometric thresholds and 95% confidence intervals are displayed as a function of test frequency in Figure 1, top panel. The CF group had significantly poorer audiometric thresholds compared to controls across all test frequencies (*p* < 0.001; See Table 2). Audiometric thresholds varied significantly across frequency, with poorer thresholds at 0.25 and 0.5 kHz and in the EHF range (14 to 16 kHz; *p* < 0.001). The interaction between Group and Test Frequency was significant (*p* < 0.001), since the mean difference between groups was not constant across all test frequencies. With regard to Race, Caucasians had significantly better audiometric thresholds compared to non-Caucasians (*p* = 0.004). Minimal mean threshold differences (± 1 dB HL) were observed from 0.25 to 8 kHz. However, at 10 and 12.5 kHz, Caucasians had mean thresholds that were 7-8 dB HL better than non-Caucasians and at 14 to 16 kHz mean thresholds were 2 to 3 dB better. Main effects of Test Ear, Sex, and Age at Test were not significant (*p* > 0.05).

**Figure 1.**
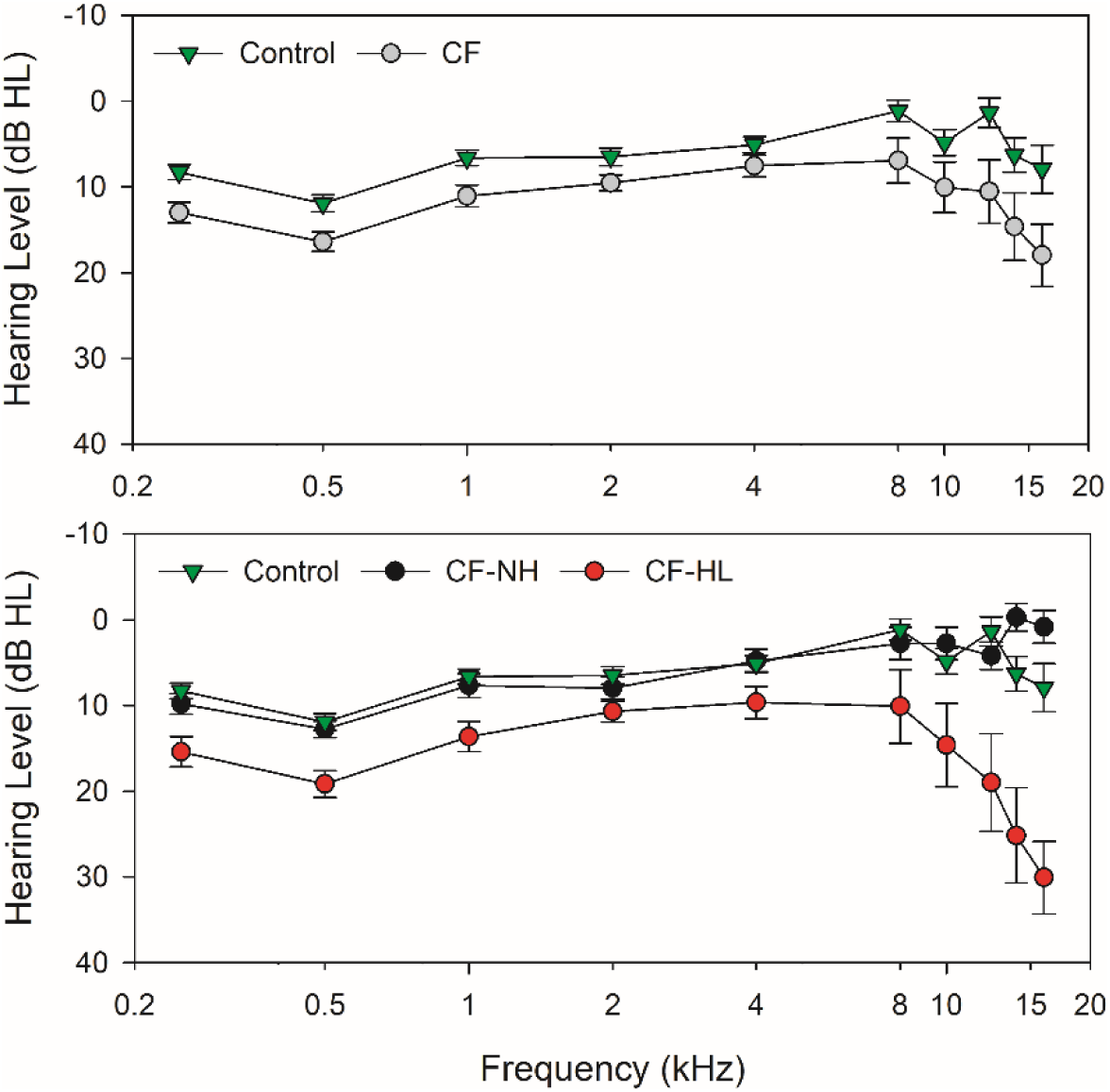
Standard and extended high frequency audiometric thresholds measured in decibel Hearing Level (dB HL) and calibrated according to ISO 389-8 and 389-1 (ISO, 2004, 2017). Top panel: Mean and 95% CI for the CF and control group. Bottom panel: Mean and 95% CI for controls with the CF group further classified into a normal hearing (CF-NH) and hearing loss (CF-HL) group.

**Table 2.** Mixed model repeated measures analysis, with p-values and F-test (Degrees of Freedom) displayed for factors included in the final model.

|  | Group | Test Frequency | Group* Frequency | Hearing Status | Group* Hearing Status | Ear | Age | Sex | Race |
| --- | --- | --- | --- | --- | --- | --- | --- | --- | --- |
| **Audiometric Threshold** | *<0.001* | *<0.001* | *0.002* | – | – | 0.541 | 0.094 | 0.349 | *0.004* |
| F (DF = 262 to 1524) | 162.18 | 37.12 | 2.94 | – | – | 0.37 | 2.81 | 0.88 | *8.50* |
| **Tympanometry** |  |  |  |  |  |  |  |  |  |
| Peak Pressure | 0.537 | – | – | *0.001*<sup>^</sup> | – | 0.593 | 0.291 | *0.002* | 0.351 |
| F (DF = 217) | 0.38 | – | – | 14.67 | – | 0.29 | 1.12 | 10.00 | 0.87 |
| Tympanometric Width | 0.156 | – | – | 0.614<sup>^</sup> | – | 0.565 | 0.175 | 0.144 | 0.088 |
| F (DF = 209 to 212) | 2.03 | – | – | 0.49 | – | 0.33 | 1.85 | 2.15 | 2.94 |
| Admittance | *0.002* | – | – | 0.127<sup>^</sup> | *0.041*<sup>^</sup> | 0.612 | 0.096 | 0.733 | 0.299 |
| F (DF = 211 to 215) | 10.35 | – | – | 2.09 | 3.23 | 0.26 | 2.80 | 0.12 | 1.09 |
| **MEMR BBN** |  |  |  |  |  |  |  |  |  |
| Ipsilateral | *0.042* | – | – | *<0.001*<sup>*</sup> | – | 0.352 | 0.051 | 0.972 | 0.392 |
| F (DF = 191) | 4.20 | – | – | 14.19 | – | 0.87 | 3.86 | *<0.00* | 0.74 |
| Contralateral | *0.004* | – | – | *0.001*<sup>*</sup> | – | 0.800 | 0.224 | 0.583 | 0.462 |
| F (DF = 183 to 185) | 8.30 | – | – | 12.42 | – | 0.06 | 1.49 | 0.30 | 0.54 |
| **Speech Perception** |  |  |  |  |  |  |  |  |  |
| SNR-Loss | *<0.001* | – | – | *0.045*<sup>*</sup> | – | 0.328 | 0.899 | *0.017* | 0.302 |
| F (DF = 181 to 182) | 23.95 | – | – | 4.07 | – | 0.96 | 0.02 | 5.83 | 1.07 |
| SRT | *<0.001* | – | – | – | – | 0.404 | 0.796 | 0.469 | 0.894 |
| F (DF = 190-191) | 30.77 | – | – | – | – | 0.70 | 0.07 | 0.53 | 0.02 |

\* Hearing status was defined as the presence of hearing loss (> 15 dB HL) at any test frequency (0.25 to 16 kHz)
<sup>^</sup> Hearing status was categorized into three groups including normal hearing, conductive or mixed hearing loss, and sensorineural hearing loss

In Figure 1, bottom panel, the CF group was separated into a normal hearing (CF-NH) and hearing loss group (CF-HL; >15 dB HL at a minimum of one audiometric test frequency), with the control group mean thresholds plotted for comparison. When the CF group was subdivided based on hearing status, the control and CF-NH group had similar thresholds from 0.25 to 16 kHz. For the CF-HL group, although thresholds are poorer across all test frequencies, the most pronounced difference was from 8 to 16 kHz where maximum thresholds ranged from 55 to 75 dB HL compared to maximum thresholds of 20 to 40 dB HL in the control group in the same frequency range.

Pearson correlation analysis was conducted to examine the relationship between participant Age at Test and average thresholds in the SF and EHF range. With all participants combined, a significant positive relationship was found between Age at Test and average EHF thresholds (*r* = .183, *p* = 0.005) but not for the average SF thresholds (*r* = .075, *p* = 0.249). Results indicate that older participants displayed higher average EHFs thresholds with no difference in SF thresholds.

### Middle Ear Function

Middle ear function was analyzed using repeated measures analysis to determine if there were differences between CF and control groups (see Table 2). A fixed effect of Hearing Status was included, where participants were categorized based on the type of hearing loss (NH, SNHL, and CHL/MHL categories). For tympanometric peak pressure, a significant effect of Group was not observed (*p* < 0.05; CF = -41.2 daPa, Control = -46.3 daPa), however Hearing Status (*p* = 0.001) and Sex (*p* = 0.002) were both significant in the model. Pairwise comparisons showed similar peak pressure for individuals with NH (mean = -19.0 daPa) and SNHL (mean = -22.4 daPa). Significantly decreased middle ear pressure was found for participants with a CHL/MHL (mean = -89.8 daPa). With regard to Sex, females (mean = -16.1 daPa) had significantly increased peak pressure compared to males (mean = -43.2 daPa).

The CF group had significantly higher admittance (mean =1.1 mmho) using 226-Hz tympanometry compared to controls (mean = 0.7 mmho; *p* = 0.002). Although Hearing Status was not significant, there was a significant Group by Hearing category interaction (*p* = 0.041). Controls had similar admittance across Hearing Status groups (NH = 0.7 mmho, CHL/MHL = 0.6 mmho, SNHL = 0.8 mmho). Within the CF group, individuals with hearing loss had higher admittance (CHL/MHL = 1.5 mmho, SNHL = 1.1 mmho) than those with normal hearing (NH = 0.8 mmho). An effect of Age at Test, Ear, or Race did not reach significance in any model for the middle ear function variables (*p* > 0.05). Additionally, for tympanometric width, there were no significant effects for any of the factors included in the model (*p* > 0.05).

### MEMR

A significant effect of Group and Hearing Status (hearing loss at any one frequency >15 dB HL) was found for both ipsilateral and contralateral MEMR thresholds. For the ipsilateral condition, the CF group had slightly lower MEMR thresholds (mean = 84.5 dB SPL) compared to controls (mean = 88.3 dB SPL; *p* = 0.042). Similarly, for the contralateral condition, individuals with CF had slightly lower MEMR thresholds (mean = 88.3 dB SPL) compared to controls (mean = 92.8 dB SPL; *p* = 0.004). Participants with hearing loss had higher MEMR thresholds than individuals with normal hearing for both the ipsilateral (NH = 83.7 dB SPL, HL = 89.1 dB SPL; *p* < 0.001) and contralateral conditions (NH = 87.5 dB SPL, HL = 92.8 dB SPL; *p* = 0.001). No significant effects were found for Test Ear, Age at Test, Sex, or Race for either ipsilateral or contralateral MEMR thresholds (*p* > 0.05). See Table 2 for repeated measures analysis results and Figure 2 for MEMR boxplots.

**Figure 2.**
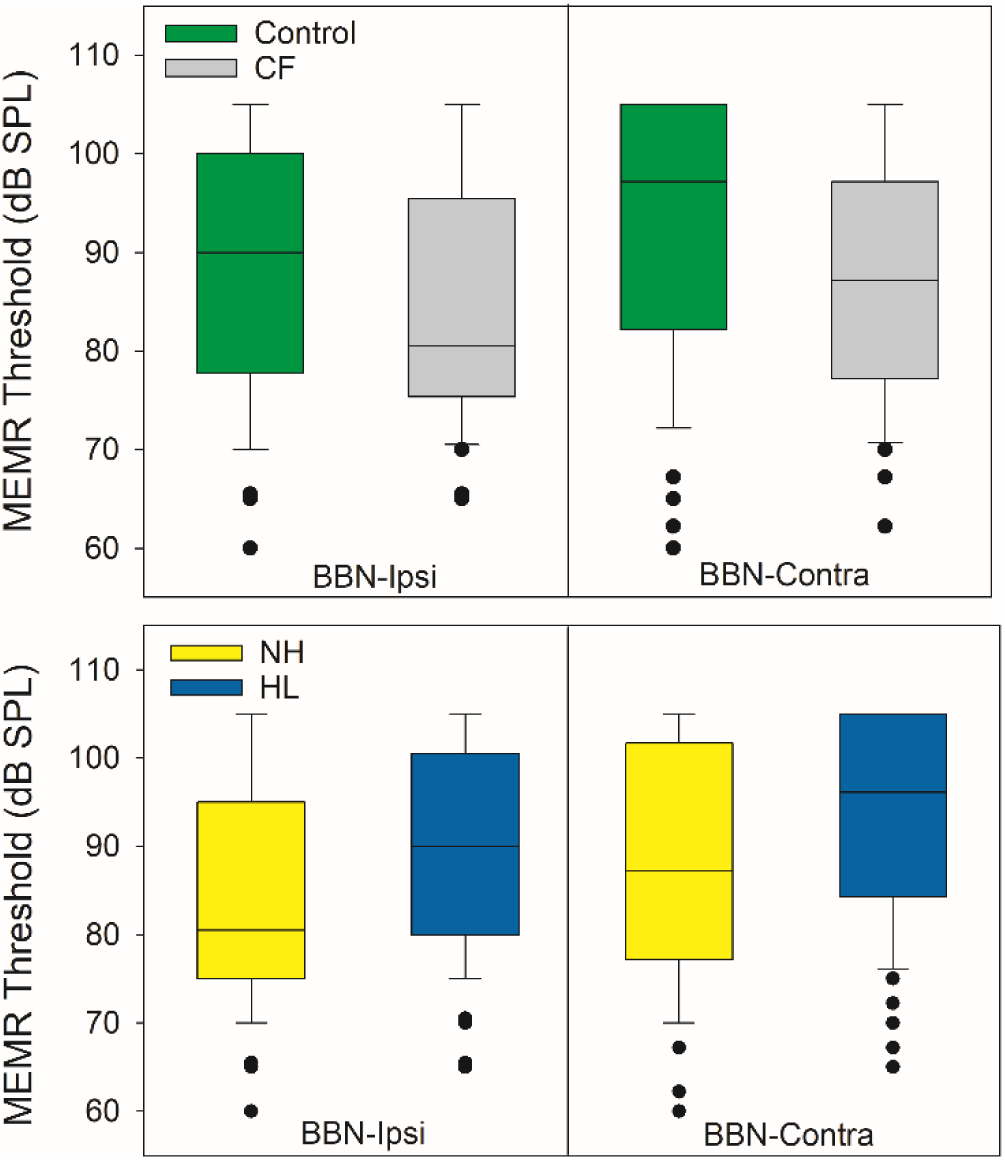
Ipsilateral and contralateral MEMR thresholds boxplots for the CF and control group (top panel) and hearing status (bottom panel). Hearing loss was defined as thresholds >15 dB HL at any test frequency (0.25-16 kHz) and included normal hearing (NH) and hearing loss (HL) categories. Boxplots show median (line), interquartile range (boxes), 95% CI (stems) and outliers (dots).

### Speech Perception

Individuals with CF showed significantly higher SRT (mean = 13.8 dB HL) compared to the control (mean = 9.5 dB HL; *p* < 0.001). In the CF group, SRTs ranged from 0 to 35 dB HL, with 81% of ears having a normal SRT and 19% with an SRT greater than 15 dB HL. In the control group, the SRT ranged from 0 to 20 dB HL with 94% of ears having a normal SRT value, and only 6% with an elevated SRT. Test Ear, Age at Test, Sex, and Race were not significant (*p* > 0.05).

For SNR-Loss (age-adjusted scores), individuals with CF had significantly poorer (higher) SNR values (mean = 2.5 dB HL) compared to controls (mean = 1.1 dB; *p* < 0.001; Figure 3, left). Within the control group, 96% of ears had normal SNR-Loss (range = -2.8 to 3.0 dB) while only 4% had a mild SNR-Loss (range = 3.5 to 4.4 dB). For the CF group, the SNR-Loss ranged from - 1.1 to 10.0 dB HL; 64% of ears had normal SNR-Loss and 36% of ears had mild or moderate SNR-Loss. In addition to a significant Group effect, individuals with HL (CF and control groups combined) had slightly higher SNR-Loss (mean = 2.2 dB) compared to those with NH (mean = 1.5 dB; *p* = 0.045; Figure 3, middle). Lastly, females had significantly higher SNR-Loss values (mean = 2.1 dB) compared to males (mean = 1.4 dB; *p* = 0.017). When the CF group was subdivided based on the presence of hearing loss, 25% of individuals with CF and normal hearing had abnormal SNR-Loss (range = 3.5 to 6 dB). In contrast, 45% of individuals with CF and hearing loss had abnormal SNR-Loss (range = 3.2 to 10 dB; Figure 3, right).

**Figure 3.**
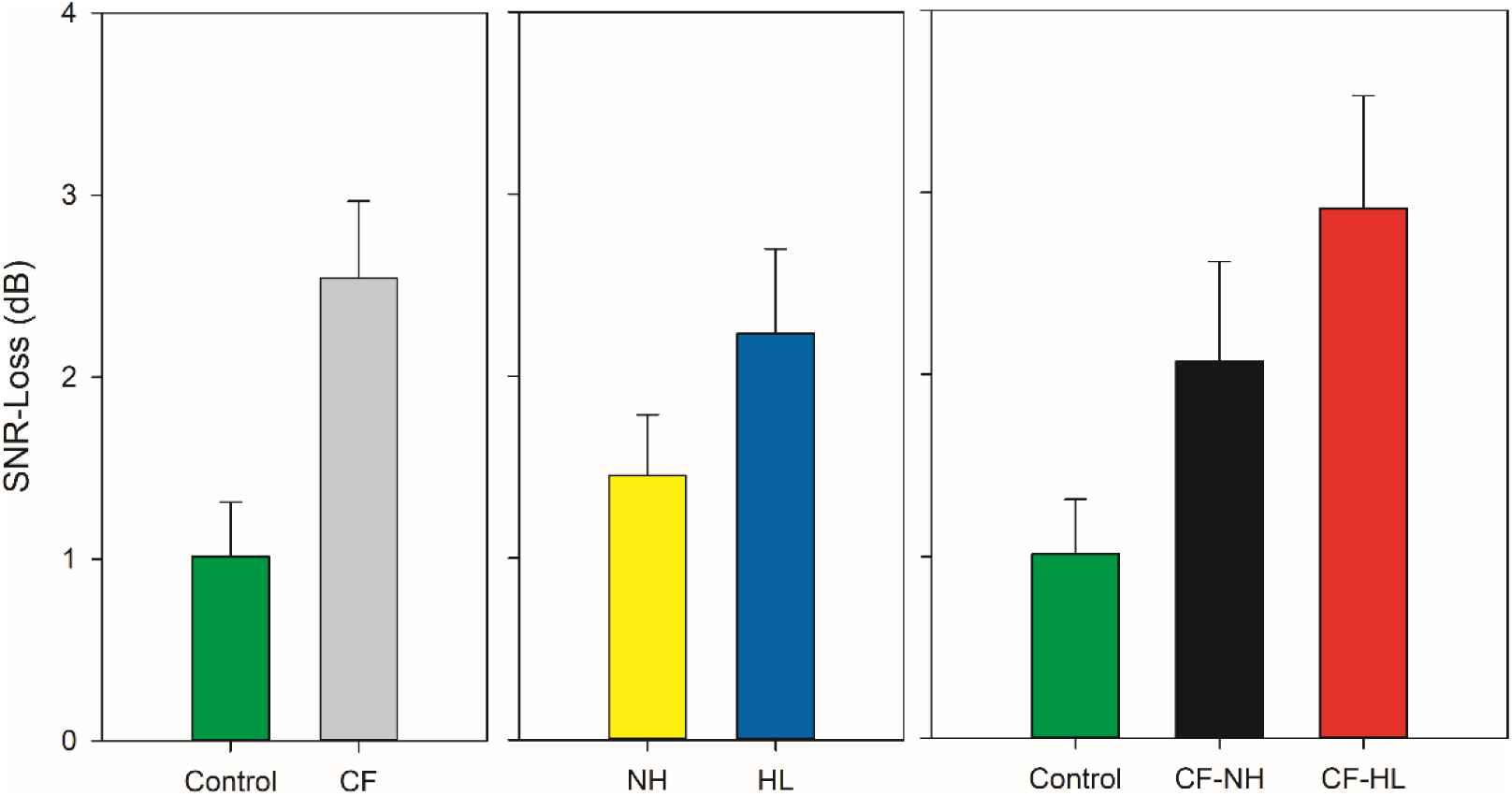
SNR-Loss displayed based on participant group (left), hearing status (middle) and lastly with the CF group further classified into a normal hearing (CF-NH) and hearing loss (CF-HL) group (right). Error bars indicate 95% confidence intervals.

To further evaluate the relationship between hearing status and speech understanding in noise, SNR-Loss was plotted against mean audiometric thresholds in the SF (0.25 to 8 kHz) and EHF (10-16 kHz) range for the CF-NH, CF-HL, and controls (see Figure 4). Correlation analysis completed with all participants combined showed a significant positive relationship for both the average SF (*r* = .301, *p* < 0.001) and EHF (*r* = .116, *p* = 0.022) thresholds compared to SNR-Loss. The SF range had a stronger relationship with SNR-Loss, explaining a greater amount of variance (9%) compared to the EHF range (3%). Lastly, upon comparison of perceived hearing concerns and the speech-in-noise performance, only 53% of the CF group with SNR-Loss reported hearing concerns on the questionnaire while 47% with an abnormal SNR-Loss reported no hearing concerns. Furthermore, only 37% of individuals with CF and hearing loss (any frequency >15 dB HL) reported hearing concerns on the questionnaire while 63% with hearing loss reported no hearing concerns.

**Figure 4.**
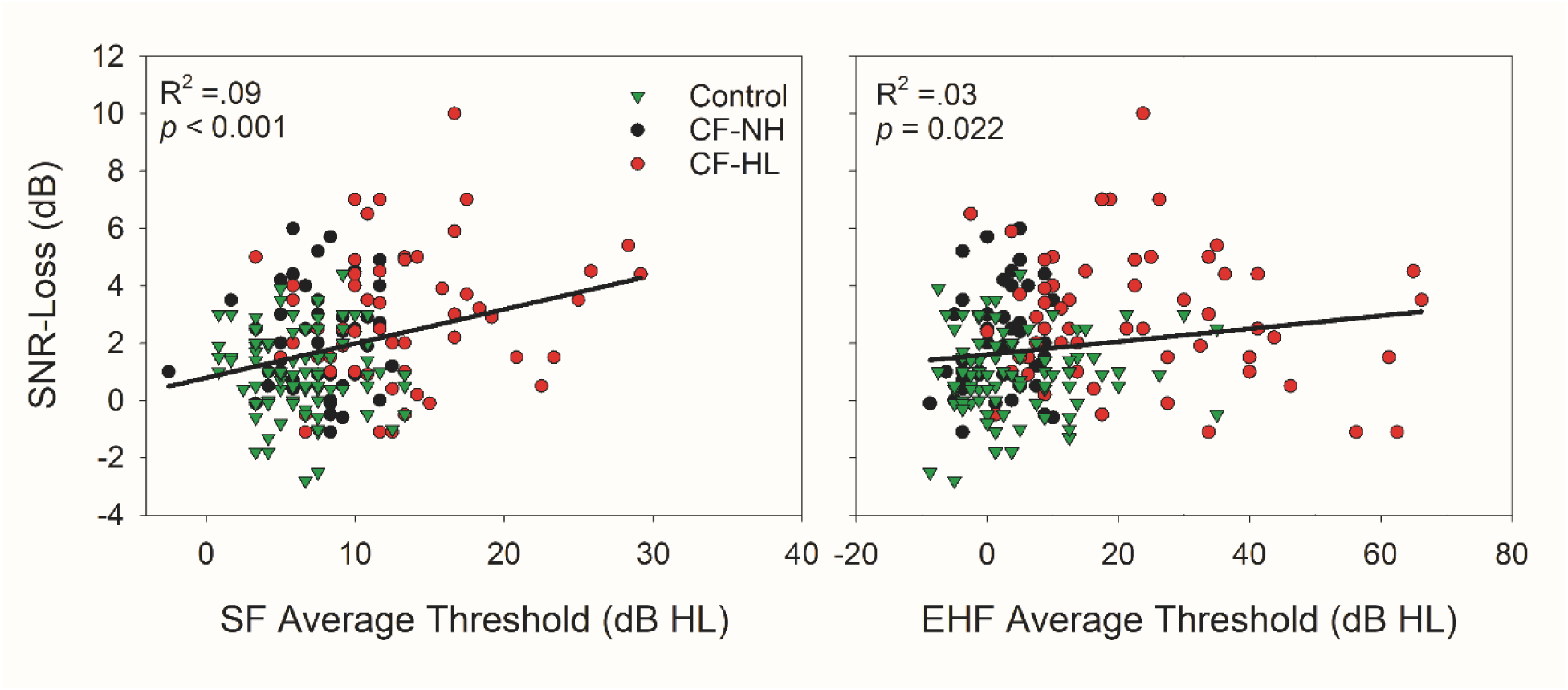
SNR-Loss plotted as a function of average hearing thresholds for the control group with the CF group further classified into a normal hearing (CF-NH) and hearing loss (CF-HL) group. Standard frequency average thresholds (SF; 0.25 to 8 kHz) are displayed on the left and extended high frequency average thresholds (EHF; 10-16 kHz) are displayed on the right. Regression lines and the coefficient of determination are displayed in each figure.

## Discussion

The main purpose of this study was to determine the prevalence of ototoxicity-related hearing loss, tinnitus, and balance and the functional impact on communication in a pediatric cohort with CF compared to age- and gender-matched controls. The CF group had a significantly higher prevalence of hearing loss in either the SF or EHF region (57%) compared to the control group (37%). The majority of controls with hearing loss, had slight-to-mild degrees of loss. In contrast, individuals with CF had high percentages of mild-to-moderate hearing loss, with some individuals displaying moderate-to-severe and severe degrees of hearing loss. The CF group also had a higher prevalence of CHL or MHL (16% combined) compared to controls (7%). Individuals with CF were also more likely to have a history of middle ear infections and PE tubes than controls. Lastly the CF group reported significantly more hearing concerns, tinnitus, balance problems than controls, had higher speech recognition thresholds in quiet, and performed poorer on a clinical speech understanding in noise task.

### Prevalence of Hearing Loss

This study showed a very high rate of hearing loss in both the standard (38%) and extended high frequency region (47%) for children, teens and young adults with CF who have received IV-AG antibiotics to treat lung exacerbations. Estimates of hearing loss vary in part due to the criterion for normal-hearing; our study included slight hearing loss (>15 dB HL) in either the SF or EHF range, so the estimates are higher than studies that have used more conservative criteria (e.g., Stavroulaki et al., 2002; Solmaz et al., 2016). Using hearing loss criteria similar to the current study, (2011)Al-Malky et al. reported clinically normal hearing (0 to 15 dB HL) from 0.25 to 12.5 kHz in children with CF (n = 45) and little to no previous exposure to IV-AGs (≤ 5 courses). However, in a high IV-AG exposure group (mean = 17.6 courses, range = 6 to 37), 35% of individuals with CF had evidence of ototoxicity in the EHF region, defined as any threshold ≥ 20 dB HL. Similarly, Al-Malky et al. (2015) reported ototoxicity in 24% of children with CF and previous exposure to IV-AGs, but in the high exposure group (range = 10 to 40 courses), 44% displayed ototoxicity with thresholds ranging from 25 to 85 dB HL.

In contrast, several studies have reported much lower rates of hearing loss, possibly due to less stringent hearing loss criteria or the exclusion of EHF audiometry. For example, Stavroulaki et al. (2002) reported normal thresholds (≤ 20 dB HL) from 0.25 to 8 kHz in a pediatric CF cohort with a history of gentamicin exposure (n = 12; range = 5.2 to 14.1 yr.) at baseline and following gentamicin treatment (4mg/kg/day; mean = 14 days; range = 11 to 29). Solmaz et al. (2016) reported 12% of individuals with CF (mean = 8.3 yr.; range = 5 to 13) with more than 3 courses of IV amikacin had hearing loss when using a pure-tone average (0.5, 1, 2, and 4 kHz) greater than 25 dB HL. However, when the hearing loss threshold was decreased to > 15 dB HL, 80% of ears in the CF group had hearing loss. In contrast, all individuals in the control group had normal hearing from 0.25 to 8 kHz by the more conservative criterion. In a large retrospective study of hearing impairment, Kreicher et al. (2018) reported 31.8% of individuals with CF (mean = 8.3 yr.) had hearing loss in at least one ear (>15 dB HL at any frequency from 0.25 to 8 kHz). Most ears with hearing loss showed a slight to mild loss (15%) with a smaller number of moderate to profound losses (3%). Martins et al. (2010) reported only 4% of a CF group (range = 0.4 to 18 yr.) displayed ototoxicity when the hearing loss criteria was based on a pure-tone average (0.5, 1, 2 kHz) >25 dB HL. However, the rate of hearing loss increased to 11% when a criterion of two or more frequencies >25 dB HL from 0.25 to 12 kHz was implemented. Lastly, Geyer, Menna Barreto, Weigert, and Teixeira (2015) reported that both the CF and control group had normal hearing in the SF region (pure-tone average of 0.25 to 8 kHz). However, in the CF group, 30.8% had at least one threshold greater than 25 dB HL in the EHF region (9-16 kHz), compared to 2.8% in the control group. When comparing thresholds across individual test frequencies, the CF group had significantly higher audiometric thresholds at 0.25, 1, 8, 9, 12.5 and 16 kHz compared to the control group (Geyer et al., 2015).

The prevalence of hearing loss in a pediatric and young adults with CF observed in the current study (57%) is comparable to studies conducted with mainly adults with CF and a higher hearing loss criterion (e.g., Garinis et al., 2017; Zettner & Folsom, 2003). Zettner and Gleser (2018) examined the progression of hearing loss in a group of 165 adults with CF and previous AG-IV exposure (mainly tobramycin) over a 10-year period. At the first audiometric examination, Zettner and Gleser (2018) reported 59% of CF group had hearing loss (≥ 25 dB HL at any frequency from 2 to 16 kHz) which progressed to 75% by their last hearing evaluation. Garinis et al. (2017) reported 56% of individuals with CF displayed hearing loss (>25 dB HL at any frequency from 0.25 to 16 kHz). Even after accounting for age and gender effects, Garinis et al. (2017) reported a significant negative effect of cumulative IV-AG doses on audiometric thresholds. Individuals with the highest IV-AG exposure (152 to 647 doses) were 4.79 times more likely to have SNHL compared to individuals with relatively low exposure (2 to 15 doses). Several other studies have also reported increased rates of hearing loss with repeated IV-AG exposure (Al-Malky et al., 2015; Al-Malky et al., 2011; Cheng et al., 2009; Mulherin et al., 1991; Pedersen et al., 1987). However, some individuals with CF and a relatively low cumulative IV-AG doses still present with significant hearing loss across the standard and extended high frequency region (Al-Malky et al., 2015; Garinis et al., 2017) indicating that a simple linear relationship between hearing and IV-AG exposure does not exist. Rather ototoxicity is the result of a complex interaction between multiple different factors, which will influence the onset and progression of hearing loss (Mulheran, Degg, Burr, Morgan, & Stableforth, 2001; Smyth, 2010). Additional risk factors that have been associated with increased ototoxicity include frequent hospitalizations and poorer lung function (Al-Malky et al., 2015; Tarshish et al., 2016), concomitant use of vancomycin (Al-Malky et al., 2015; O’Donnell et al., 2010), diabetes (Kreicher et al., 2018), mitochondrial mutations, (Al-Malky et al., 2015), and Hispanic or Asian ethnicity (Knight, Kraemer, & Neuwelt, 2005; O’Donnell et al., 2010).

### Importance of EHFs in Ototoxicity Monitoring

It is well established that aminoglycoside induced hearing loss originates in the EHF region and with increased IV-AG exposure, eventually progresses to the SF region (Fausti et al., 2003; Fausti et al., 1999). Therefore, the inclusion of EHF audiometry when monitoring for ototoxicity is essential. Several studies have evaluated the presence of hearing loss based on frequency region. For example, Al-Malky et al. (2015) showed that the use of EHF (9-16 kHz) audiometry identified 15 children with hearing loss, while SF audiometry only detected 13 children. When using a SF pure-tone average (0.5, 1, 2 kHz), Martins et al. (2010) reported only 4% of CF cases had hearing loss (> 25 dB HL), however when EHFs (9-12 kHz) were included, 11% had hearing loss. Geyer et al. (2015) reported that while all individuals with CF had normal hearing in the SFs, 30.8% of the CF group had EHF hearing loss (9-16 kHz). Furthermore, Garinis et al. (2017) identified 45 individuals with CF with hearing loss (>25 dB HL at any frequency from 0.25 to 16 kHz), of which 28 participants only displayed hearing loss at frequencies > 9 kHz. Hearing loss in the aforementioned studies is presumed to be sensorineural since the studies excluded individuals with history of chronic middle ear infections, abnormal middle ear status, middle ear effusion, tympanic membrane perforation, or conductive hearing loss verified with BC thresholds. In the current study, 38% of the CF group had hearing loss in the SF and 47% had hearing loss in the EHF region. When examining audiometric thresholds across frequency region, 15 individuals with CF (n = 22 ears, 7 bilateral, 8 unilateral) had normal hearing in the SFs but displayed either slight to mild (n = 16 ears) or moderate to moderately-severe (n = 6 ears) hearing loss in the EHFs. Therefore, the use of only SF audiometry would result in the misdiagnosis of 15 individuals with CF with normal hearing, when they display evidence of significant ototoxicity in the EHFs. Furthermore, older participants (CF and controls grouped together) displayed higher average EHF thresholds, with no effect of age on average SF thresholds. Since individuals in the study are still very young, this may suggest more treatment exposure for individuals with CF when the groups are separated out.

### Conductive Hearing Loss and History of Otitis Media

The mucosal epithelium of the middle ear and eustachian tube is contiguous with the upper respiratory track. Thus, it was previously assumed that individuals with CF may have a higher that usual incidence of middle ear disease. However, temporal bone studies of individuals with and without CF have shown similar pneumatization and mucosal histology of the middle ear (Berkhout et al., 2014; Seifert et al., 2010; Todd & Martin, 1988; Yildirim et al., 2000). In addition, several studies have suggested that individuals with CF are at no higher risk than age-matched controls, with some studies showing lower rates of inflammatory ear disease compared to controls (Bak-Pedersen & Larsen, 1979; Cepero et al., 1987; Cipolli et al., 1993; Forcucci & Stark, 1972; Jorissen, De Boeck, & Feenstra, 1998; Martins et al., 2011). However, most previous studies did not include bone conduction audiometry, and if tympanometry was included, individuals with middle ear issues were excluded (otitis media, tympanic membrane perforation). Therefore, most studies were not designed to detect conductive hearing loss or middle ear disorders, which may help to explain why this issue has gone unrecognized.

Previously in the CF literature, the prevalence of otitis media have ranged from 3 to 45%. A retrospective analysis of 450 individuals with CF revealed only 3% had a history of otologic disease with only 1% receiving PE tubes (Cepero et al., 1987). In a CF group (n = 80), Forman-Franco et al. (1979) reported 24% had a history of acute otitis media and only one individual had received PE tubes. Bak-Pedersen and Larsen (1979) reported slightly higher rates of a history of acute otitis media (35%) in a cohort of 111 individuals with CF. In a retrospective pediatric and adult cohort study, Cheng et al. (2009) reported 20% had a history of middle ear effusion and or eustachian tube dysfunction and abnormal tympanometry (Type B and C). Most recently, Kreicher et al. (2018) conducted a retrospective study of 217 children with CF and found 45% had acute otitis media and 29% were diagnosed with chronic otitis media. The number of otitis media infections across patients ranged from 1 to 66, with an average of 6.6 per patient. PE tubes were placed in 8% of the CF group and 23% were diagnosed with Eustachian tube dysfunction. Of the 94 ears with tympanometry results, 19% had abnormal tympanograms (type B and C).

With regard to the prevalence of CHL in individuals with CF, previous studies have reported relatively low percentages ranging from 0 to 10% (Al-Malky et al., 2015; Al-Malky et al., 2011; Geyer et al., 2015; Martins et al., 2010). In a group of children and adults with CF (n = 80) with previous tobramycin and gentamicin exposure, Forman-Franco et al. (1979) reported no individuals displayed CHL. Pedersen et al. (1987) reported a slightly higher rate, with 7% of individuals with CF diagnosed with a CHL. Using BC thresholds measured from 0.25 to 4 kHz, Kreicher et al. (2018) reported that 10% had a conductive component to their hearing loss, with equal percentages of conductive (5.3%) and mixed (5.1%) loss. Lastly in a cohort of 70 children with CF, Kulczycki, Butler, McCord-Dickman, and Herer (1970) reported a much higher rate with 27% of study participants showing a mild conductive loss.

In the present study, 118 participants (CF = 57, Control = 61) were investigated with the use of the traditional approach of audiometric and otologic verification to detect and identify ear disease. In addition, 226-Hz tympanometry and MEMRs were performed. The physiologic data were used to analyze the pressure in the middle ear space, integrity and mobility of the tympanic membrane and ossicles, and stapedius muscle function. We found that 42% of participants with CF reported a history of chronic otitis media, 16% have had PE tube surgery, and 16% had a conductive component to their hearing loss, compared to 26%, 11% and 7% in the control group, respectively. Thus, compared to an age and gender matched sample, our CF group had significantly more reports of past otitis media with effusion and PE tube placement. The occurrence of CHL in our sample was mostly mild and was not accompanied by current otitis media with effusion or flat tympanometry. Rather, in most cases, tympanometry showed significantly more negative peak pressure in the ears with CHL, indicating Eustachian tube dysfunction. Most cases of CHL also showed abnormally high admittance for tympanometry, although this was not statistically significant in terms of hearing loss type overall. Histories of PE tubes are frequently associated with high admittance due to tympanic membrane defects related to either spontaneous perforation or myringotomy of the tympanic membrane (Hunter & Blankenship, 2017).

The finding of lower (better) MEMR thresholds in individuals with CF and AG exposure for both ipsilateral and contralateral stimuli has not been reported previously and does not have a clear explanation (However, see Westman et al., 2020 in this volume).While MEMR thresholds in the present study were overall slightly lower in the CF group, MEMR thresholds were poorer for individuals with hearing loss. This result was expected and is consistent with changes in MEMRs due to outer hair cell loss in kanamycin exposure (Borg & Engström, 1982). Validation in another sample, and with non-AG exposed participants is needed to explore the finding of better MEMR in patients with normal hearing.

### Functional Impact

While most previous studies have focused on audiometric assessment it is important to evaluate the functional impact of the hearing loss on the individual and their communication abilities. In the current study, 32% of individuals with CF reported hearing difficulties and 53% reported tinnitus compared to 8% and 16% in the control group, respectively. In comparison, Al-Malky et al. (2015) reported that most children with ototoxicity did not report any issues with hearing or tinnitus. Of the 15 individuals with CF that displayed hearing loss, only four reported issues with both hearing and tinnitus, two had hearing difficulties, and one child reported issues with tinnitus. The remaining 8 children with ototoxicity did not report any issues with hearing or tinnitus. Similarly, Mulheran et al. (2001) reported 17% of the CF group had significant ototoxicity, many with hearing loss that had progressed into the SF region. Yet most of these individuals were asymptomatic; none of the patients had self-report of hearing difficulties and only four individuals experienced periodic tinnitus during treatment. Clinical measures of speech understanding in quiet and noise are important tools to evaluate the impact of hearing impairment on communication abilities. The CF group had significantly poorer speech recognition thresholds and speech-in-noise scores compared to age-matched controls. For speech understanding in quiet, 6% of controls had an abnormal SRT (>15 dB HL) compared to 19% of the CF group. On the BKB-SIN, 64% of the CF group had an abnormal SNR-Loss that ranged from mild to moderate, yet only 4% of the controls had an abnormal SNR-Loss, all within the mild range. While 45% of individuals with CF with hearing loss also had an abnormal SNR-Loss, 25% of individuals with CF and normal hearing displayed an abnormal SNR-Loss. Although several studies have reported that speech perception was conducted as part of the audiometric test battery, test results were not provided/reported (Forman-Franco et al., 1979; Martins et al., 2010; Pedersen et al., 1987). Forcucci and Stark (1972) conducted audiometric, ototologic and speech-language examinations in a group of 31 children with CF and 22% had deficient speech-language development. Due to the cumulative ototoxic nature of IV-AG, individuals with CF are at a higher risk for developing hearing loss that can impact speech and language development, literacy development, scholastic achievement, and overall quality of life (Moeller et al., 2007; Yoshinaga-Itano, 1999, 2003), (Moeller et al., 2007). Therefore, ototoxicity monitoring/management programs should include functional impact measures that can be administered quickly, ideally at the patient bedside or at home. Even if the AG treatment schedule cannot be altered to minimize ototoxicity, self-report measures and clinical speech perception assessments are necessary to evaluate the impact of hearing loss and to identify individuals for audiologic treatment. Additionally, there is a substantial lack of patient-reported outcomes in ototoxicity monitoring programs which are critical for patient-centered clinical care.

Very few studies have reported the effects of AG treatment on vestibular function in individuals with CF (Handelsman et al., 2017; Scheenstra, Rijntjes, Tavy, Kingma, & Heijerman, 2009). Furthermore, studies report wide variability in presence of vestibular impairments and very little has been reported about vestibular asymmetries. Scheenstra et al. (2009) reported 35% of adults with CF had either a peripheral or central impairment as diagnosed using Electronystagmography with caloric irrigation, while only 16% self-reported vestibular issues on a questionnaire. In a group of children and adults with CF, Handelsman et al. (2017) reported that while most underreported symptoms, 79% had vestibular system dysfunction based on diagnostic test battery (dynamic visual acuity, videonystagmography, sinusoidal and rotational step testing). In the current study 32% of the CF group reported balance issues compared to 16% in controls. The presence of vestibular impairments is high among individuals with CF, yet most go undiagnosed due to lack of testing and underreport of patient symptoms. Vestibular assessments that can be performed quickly with minimal physical or psychological discomfort, such as the video head impulse test and vestibular evoked myogenic potential test, should be included in ototoxicity monitoring programs.

### Limitations and Future Directions

The primary limitation of this study is that individuals with CF had pre-existing exposure to AGs, and thus nearly half had pre-existing hearing loss as well. Efforts are underway to continue longitudinal assessment of this sample, and to expand the assessment to children not yet exposed to ascertain onset and progression of hearing loss in future studies. In process for this study is analysis of past doses of AGs, and pharmacodynamic measures (peak, trough blood levels and deeper tissue estimation of drug exposure over time) to determine relationships to hearing loss and changes over time with repeated hearing tests. While the sample size is relatively modest, it is one of the largest studies published specifically in children with prospective assessment. Diabetes and genetic variants that are related to AG ototoxicity as well as known mutations (Ex: m1555A>G mutation) that increase risk for developing hearing loss will be included in future studies.

## Conclusion

Individuals with CF are at extremely high risk for developing hearing loss and balance disorders due to routine exposure of IV-AGs to treat pulmonary exacerbations. As the predicted median age of survival for individuals with CF continues to increase, many will become exposed to very high cumulative doses of IV-AG, thereby increasing their risk for developing hearing loss. Our results showed high rates of SNHL and CHL in the CF group, higher than most pediatric CF studies due to the inclusion of EHF audiometry and a more stringent hearing loss criteria (threshold at any frequency >15 dB HL). Furthermore, older participants display higher average EHF thresholds, with no effect of age on average SF thresholds. Studies that based the presence of hearing loss on the individual thresholds within the SF region, a SF pure-tone average, or higher significant threshold criteria (dB HL) are likely underestimating the incidence of hearing loss in this population. Furthermore, these individuals with CF displayed high rates of middle ear dysfunction and conductive hearing loss, a problem that has not been identified in studies that did not include bone conduction or excluded individuals with abnormal middle ear status.

Previous studies have not reported speech perception in either quiet or noise. Our study shows that SNHL connected with AG use degrades the listener’s ability to perceive speech-in-noise which may diminish quality of life and has clear educational implications. The impacts of broadened tuning resulting from EHF hearing loss on speech-in-noise are just beginning to be appreciated (Besser et al., 2015; Hunter et al., 2020b), but have received scant attention in patients with ototoxicity. The importance of speech communication for patients with chronic illness also has implications for ongoing care. Medical staff must wear masks due to contact isolation requirements, and they report that communication is challenging with their patients with hearing loss. Armed with evidence of ototoxicity, physicians report that they are willing to consider treatment modifications to minimize permanent hearing loss and the impact on communication (Garinis et al., 2018). The BKB-SIN test used in this study to evaluate speech-in-noise performance are derived from child language samples and the sentences are at approximately a first-grade reading level (Nilsson, Soli, & Sullivan, 1994). The test is adaptive, avoiding ceiling and floor effects, has ecological validity and is reportedly not confounded by language abilities and working memory (Magimairaj, Nagaraj, & Benafield, 2018). However, other studies have found that sentence in noise performance is associated with language and working memory in children (McCreery, Spratford, Kirby, & Brennan, 2017). The BKB-SIN test yields a SNR-Loss score that is age-normalized, allowing results to be compared across different populations. Inclusion of an age- and language-appropriate speech-in-noise measure is recommended both to evaluate functional effects of ototoxicity, and to assess the need for, and benefit of amplification.

Lastly, these the CF group reported significantly more issues with hearing (32%), tinnitus (53%), and balance (32%), compared to controls (8%, 16%, 9%), respectively. However, self-report of hearing difficulties was not predictive of hearing loss or speech-in-noise performance, thus self-report cannot replace formalized testing to monitor for evidence of ototoxicity.

## Abbreviations

AC: Air-Conduction
AG: Aminoglycoside
BKB-SIN: Bamford-Kowal-Bench Speech-in-Noise
BC: Bone-conduction
CCHMC: Cincinnati Children’s Hospital Medical Center
CHL: Conductive Hearing Loss
CF: Cystic Fibrosis
EHF: Extended High Frequency
IV-AG: Intravenous-Aminoglycoside
MHL: Mixed Hearing Loss
Normal Hearing: NH
Pressure Equalization: PE
Signal-to-Noise Ratio: SNR
Sensitive (Frequency) Range for Ototoxicity: SRO
Sensorineural Hearing Loss: SNHL
Speech Reception Threshold: SRT
Standard Frequency: SF

## Data Availability

The data that support the findings of this study are available from the corresponding author, upon reasonable request.

## Acknowledgments

This research was supported by the National Institutes of Health, National Institute on Deafness and Other Communication Disorders Grant R01DC010202 (multi-principal investigators: Doug Keefe, Lisa L. Hunter and Patrick Feeney), National Institutes of Health, Clinical and Translational Science Award Program Grant 5UL1TR001425-04 (Center for Clinical and Translational Science Training at the University of Cincinnati) and the Cincinnati Children’s Hospital Medical Center Research Foundation Place Outcomes Research Award (multi-principle investigators: Lisa L. Hunter and John P. Clancy). The content of this manuscript is solely the responsibility of the authors and does not reflect the official views of the National Institute of Health or Cincinnati Children’s Hospital Medical Center. Portions of this study were presented at the World Congress of Audiology 2018, North American Cystic Fibrosis Conference 2019, and the Association for Research in Otolaryngology 2020. We also thank our participants, their families, and Summer Undergraduate Research Foundation (SURF) scholars.

